# From Hospital to Home: Continuity between Skin-to-Skin Care and Later Verbal Engagement in Infants Born Preterm

**DOI:** 10.1101/2025.07.02.25330762

**Authors:** Pamela M. Rios, Virginia A. Marchman, Molly F. Lazarus, Nuria L. Ontiveros Perez, Melissa Scala, Katherine E. Travis, Heidi M. Feldman

## Abstract

This descriptive cohort study documented continuity between family-delivered skin-to-skin care rates for preterm infants in the NICU and amount of child-directed speech at child age 9 months. Involvement in skin-to-skin care may be an early marker of caregiver engagement and a target for early interventions that support positive caregiver-infant interactions.

---

Children born very preterm (VPT; <32 weeks gestational age (GA)) are at elevated risk for neurodevelopmental delays.^1,2^ Skin-to-skin (STS) care, in which caregivers hold their infant against their bare chest, is a developmental care practice linked to improved outcomes. Specifically, higher rates of family-administered STS care in the Neonatal Intensive Care Unit (NICU) have been associated with favorable neurodevelopmental outcomes at 12 and 18 months (all ages adjusted for the degree of prematurity).^3,4^ One possible interpretation is that associations reflect direct, long-term benefits of developmental care on neurodevelopment. A second possibility is that STS care is linked to later caregiver engagement that directly or cumulatively leads to positive neurodevelopmental outcomes. Exploring these possibilities requires data on how STS care in the hospital relates to caregiver engagement at home.

The current descriptive cohort study of VPT infants aimed to investigate relations between rates of family-administered STS care in the NICU and caregiver engagement in the home environment in infancy. In infancy and toddlerhood, one index of caregiver engagement is the quantity of child-directed speech, which has been linked to neurodevelopmental outcomes in full term and preterm infants.^5^ Assessing continuity in caregiver engagement from hospital to home may help explain how early experiences contribute to neurodevelopmental outcomes. In addition, rates of STS care may serve as an early marker of caregiver engagement, indicating families who may benefit from early interventions that support positive caregiver-infant interactions.

## Methods

### Participants

Participants (*n* = 31, 54.8% female) were infants born VPT, recruited between June 2021 and May 2024 from Lucile Packard Children’s Hospital (LPCH) in Stanford and California’s High Risk Infant Follow-Up Clinic (HRIF). Exclusion criteria were congenital or genetic anomalies, hearing or vision loss, caregiver primary language other than English or Spanish, and missing developmental care or caregiver verbal engagement data. This study was reviewed and approved by the Stanford School of Medicine Institutional Review Board (IRB; Protocol #53904 and #44480), and informed consent was obtained for each participant.

### Measures

#### Clinical and Demographic Information

Demographic and clinical characteristics of infants, including GA (weeks), length of hospital stay (days), and child sex (male or female), were extracted from the electronic medical record (EMR). Race, ethnicity, and language spoken at home were reported by caregivers at the time of enrollment. Socioeconomic status (SES) was measured using the Hollingshead Four Factor Index^6^, which combines caregivers’ education and occupation into a composite score (range = 8 to 66). Clinical data regarding four common comorbidities of premature birth were also obtained from the EMR. Bronchopulmonary dysplasia (BPD) was defined by grade two or higher severity, intraventricular hemorrhage (IVH) by any grade present, sepsis by positive blood culture or > seven days of antibiotics, and necrotizing enterocolitis by medical or surgical diagnosis. A binary health acuity variable was derived, classifying infants as having either none (health acuity = 0) or one or more (health acuity = 1) of these conditions.

#### Family-Involved Visitation and Developmental Care Practices

Information regarding visitation and developmental care practices were extracted from the EMR. Nurses at the LPCH NICU routinely document all instances of visitation to bedside by family members. To account for varying lengths of hospitalization, visitation frequency was computed as the total number of visitation instances divided by the number of days of hospital stay. Nurses also routinely record all instances of STS care, the duration of each session in minutes, and the individual(s) involved (e.g., mother, father, staff member). For the current study, only STS care instances in which one or more family members were involved were analyzed. Rates of family-delivered STS care were computed as the sum of the total number of minutes of STS, divided by the number of days of hospital stay.

#### Caregiver Verbal Engagement

Caregiver verbal engagement was assessed when the child was approximately 9 months using day-long audio recordings via the Language Environment Analyses (LENA) system.^7^ The LENA system consists of a high-quality digital recording device worn by the infant in specialized clothing. Families were instructed to begin recording during a typical day at home and allow it to run automatically for up to 16 hours. The LENA speech-recognition software classifies all adult speech that is “near and clear” to the child and generates adult word counts (AWC) for each 5-minute segment of the recording. All recordings were > 4 hours. Recordings were subsequently cleaned using an automated classifier to identify 5-minute segments during which the child was not likely to be sleeping and that were most likely to contain child-directed speech (CDS).^8^ Child-directed adult word counts/hour (CD-AWC/hour) were computed as the sum of AWC during those non-sleep, CDS segments, divided by the length of the cleaned recording.

#### Statistical Analyses

All analyses were conducted in R version 4.4.3. Both STS care rate and CD-AWC/hour were inspected for normality using histograms and Shapiro-Wilks tests. STS care rate was significantly skewed, so it was transformed using a base 10 log. Hierarchical linear regression models were used to examine relations between STS care rates and later CD-AWC/hour. SES and health acuity were selected *a priori* as covariates, given their anticipated relevance to STS care rates and verbal engagement measures. Family visitation frequency was included as a covariate to assess the specific effects of engaging in STS care activities beyond hospital visitation alone.

## Results

Table 1 summarizes the clinical and demographic characteristics of the sample. All infants were born VPT (<32 weeks GA), with a mean GA of about 29 weeks. The average length of hospital stay was around 73 days. Table 2 outlines NICU visitation frequency, STS care rates, day-long recording lengths, and CD-AWC/hour. On average, families visited around once a day, with some families visiting as often as 2-3 times/day. Infants experienced STS for about 24 mins/day. However, there was substantial variation, such that one infant received more than 100 mins/day, whereas two infants received none. At child age 9 months, portions of the day-long recordings that were likely to contain child-directed speech averaged about 8 hours. Caregiver verbal engagement varied widely across families, such that one infant heard more than 3500 words/hour, whereas another infant heard only 426 words/hour.

**Table 1.**
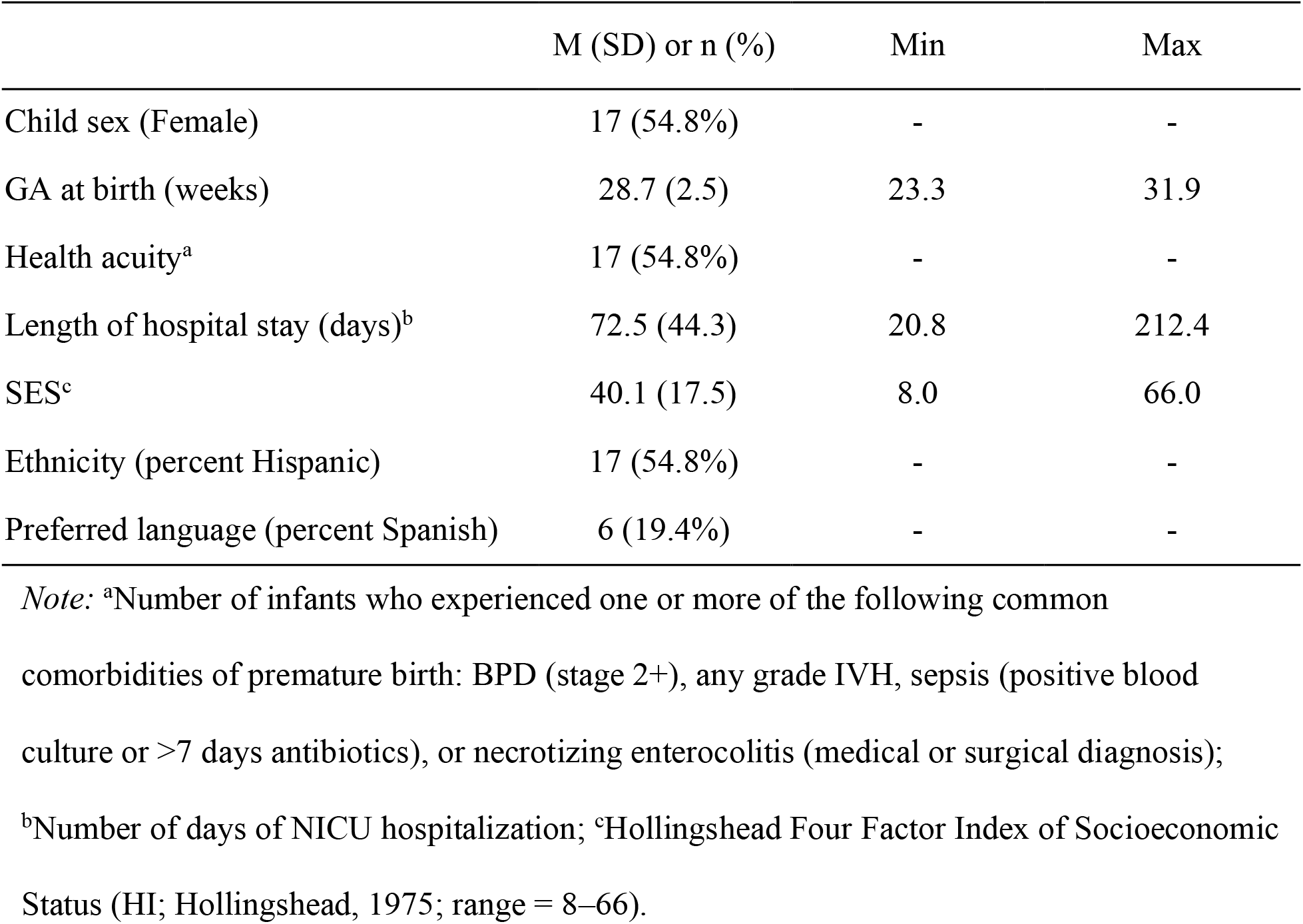
Descriptive statistics for clinical variables of all infants (*n* = 31).

|  | M (SD) or n (%) | Min | Max |
| --- | --- | --- | --- |
| Child sex (Female) | 17 (54.8%) | - | - |
| GA at birth (weeks) | 28.7 (2.5) | 23.3 | 31.9 |
| Health acuity<sup>a</sup> | 17 (54.8%) | - | - |
| Length of hospital stay (days)<sup>b</sup> | 72.5 (44.3) | 20.8 | 212.4 |
| SES<sup>c</sup> | 40.1 (17.5) | 8.0 | 66.0 |
| Ethnicity (percent Hispanic) | 17 (54.8%) | - | - |
| Preferred language (percent Spanish) | 6 (19.4%) | - | - |

*Note:* <sup>a</sup>Number of infants who experienced one or more of the following common comorbidities of premature birth: BPD (stage 2+), any grade IVH, sepsis (positive blood culture or >7 days antibiotics), or necrotizing enterocolitis (medical or surgical diagnosis); <sup>b</sup>Number of days of NICU hospitalization; <sup>c</sup>Hollingshead Four Factor Index of Socioeconomic Status (HI; Hollingshead, 1975; range = 8–66).

**Table 2.** Descriptive statistics for measures in the NICU and day-long home recordings (*n* = 31).

|  | M (SD) | Min | Max |
| --- | --- | --- | --- |
| Visitation frequency (visits/day)<sup>a</sup> | 1.3 (0.5) | 0.2 | 2.6 |
| Skin-to-skin care rate<sup>b</sup> | 24.4 (26.8) | 0.0 | 107.8 |
| Child age at day-long recording<sup>c</sup> | 9.7 (0.8) | 8.6 | 11.9 |
| Duration of total day-long recording<sup>c</sup> | 15.3 (2.2) | 4.6 | 16.0 |
| Duration of CDS in day-long recording<sup>c</sup> | 7.8 (2.5) | 2.4 | 14.6 |
| CD-AWC/hour<sup>d</sup> | 1787.2 (784.6) | 425.8 | 3565.0 |

*Note:* <sup>a</sup>Number of family visits to bedside/day of NICU hospitalization; <sup>b</sup>Minutes of family-involved skin-to-skin care/day of NICU hospitalization; <sup>c</sup>Child adjusted age, total recording length, and CDS recording length of day-long recordings; <sup>d</sup>Number of child-directed adult words/hour during day-long recording.

We next examined whether infants who received more STS care in the NICU were also exposed to greater verbal engagement at 9 months. Table 3 shows that a model with SES, health acuity, and visitation frequency (Model 1) accounted for approximately 13% of the variance in CD-AWC/hour. Model 2 shows that variation in STS care significantly increased the overall prediction of CD-AWC/hour (*F*(1, 26) = 7.13, *p* = 0.01), accounting for approximately 19% additional variance. As Figure 1 illustrates, caregivers who provided more STS care in the NICU also provided more CD-AWC/hour when the child was 9 months. For example, increasing STS care rate from 25 to 50 minutes/day (a 0.29 increase) would be associated with an increase of 186 child-directed adult words/hour.

**Table 3.**
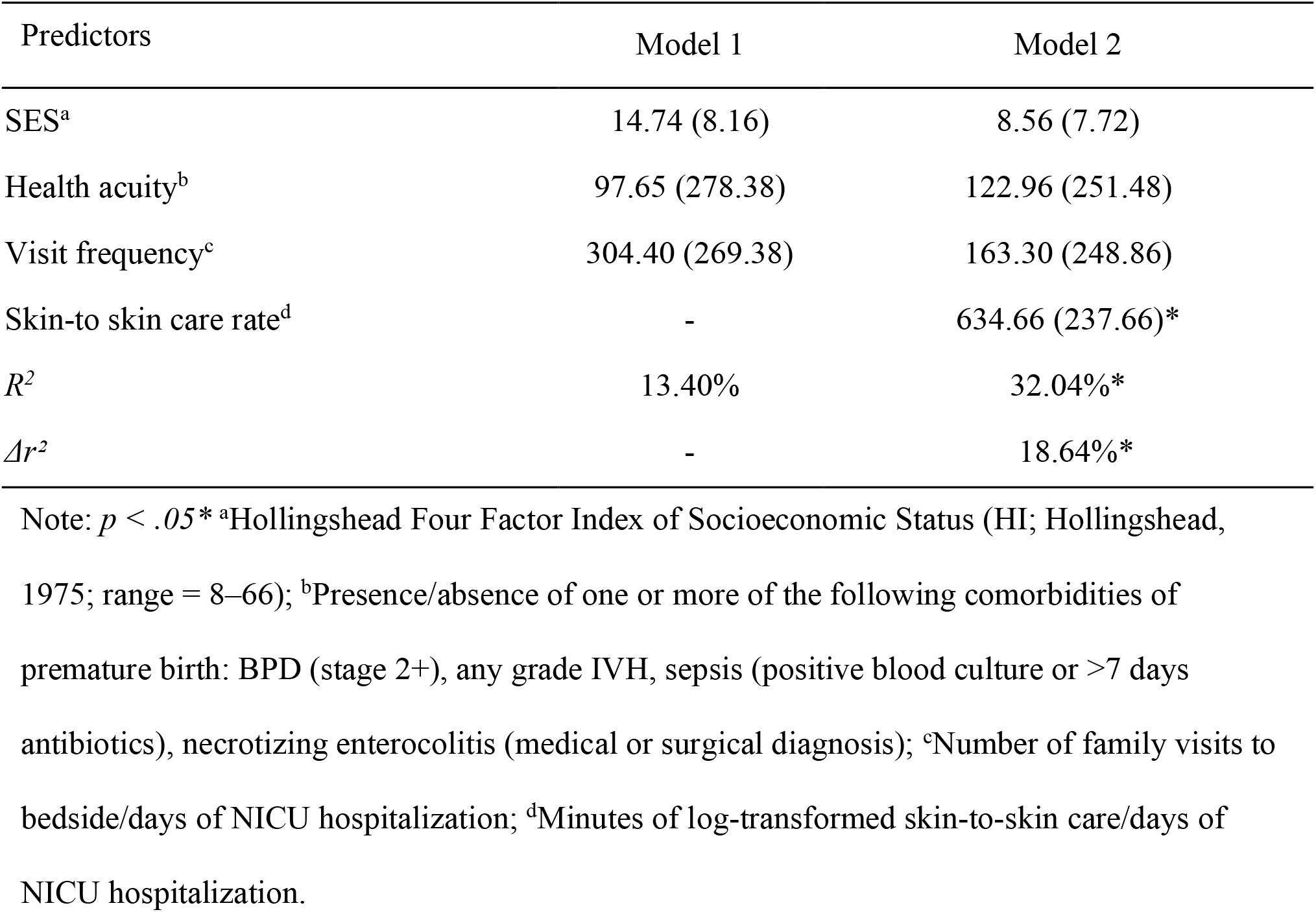
Multiple regression models (unstandardized coefficients (SE)) predicting child-directed adult word counts/hour at 9-months adjusted age, after consideration of covariates (*n* = 31).

| Predictors | Model 1 | Model 2 |
| --- | --- | --- |
| SES<sup>a</sup> | 14.74 (8.16) | 8.56 (7.72) |
| Health acuity<sup>b</sup> | 97.65 (278.38) | 122.96 (251.48) |
| Visit frequency<sup>c</sup> | 304.40 (269.38) | 163.30 (248.86) |
| Skin-to skin care rate<sup>d</sup> | - | 634.66 (237.66)* |
| $R^2$ | 13.40% | 32.04%* |
| $\Delta R^2$ | - | 18.64%* |

Note: $p < .05$ \* <sup>a</sup>Hollingshead Four Factor Index of Socioeconomic Status (HI; Hollingshead, 1975; range = 8–66); <sup>b</sup>Presence/absence of one or more of the following comorbidities of premature birth: BPD (stage 2+), any grade IVH, sepsis (positive blood culture or >7 days antibiotics), necrotizing enterocolitis (medical or surgical diagnosis); <sup>c</sup>Number of family visits to bedside/days of NICU hospitalization; <sup>d</sup>Minutes of log-transformed skin-to-skin care/days of NICU hospitalization.

**Figure 1.**
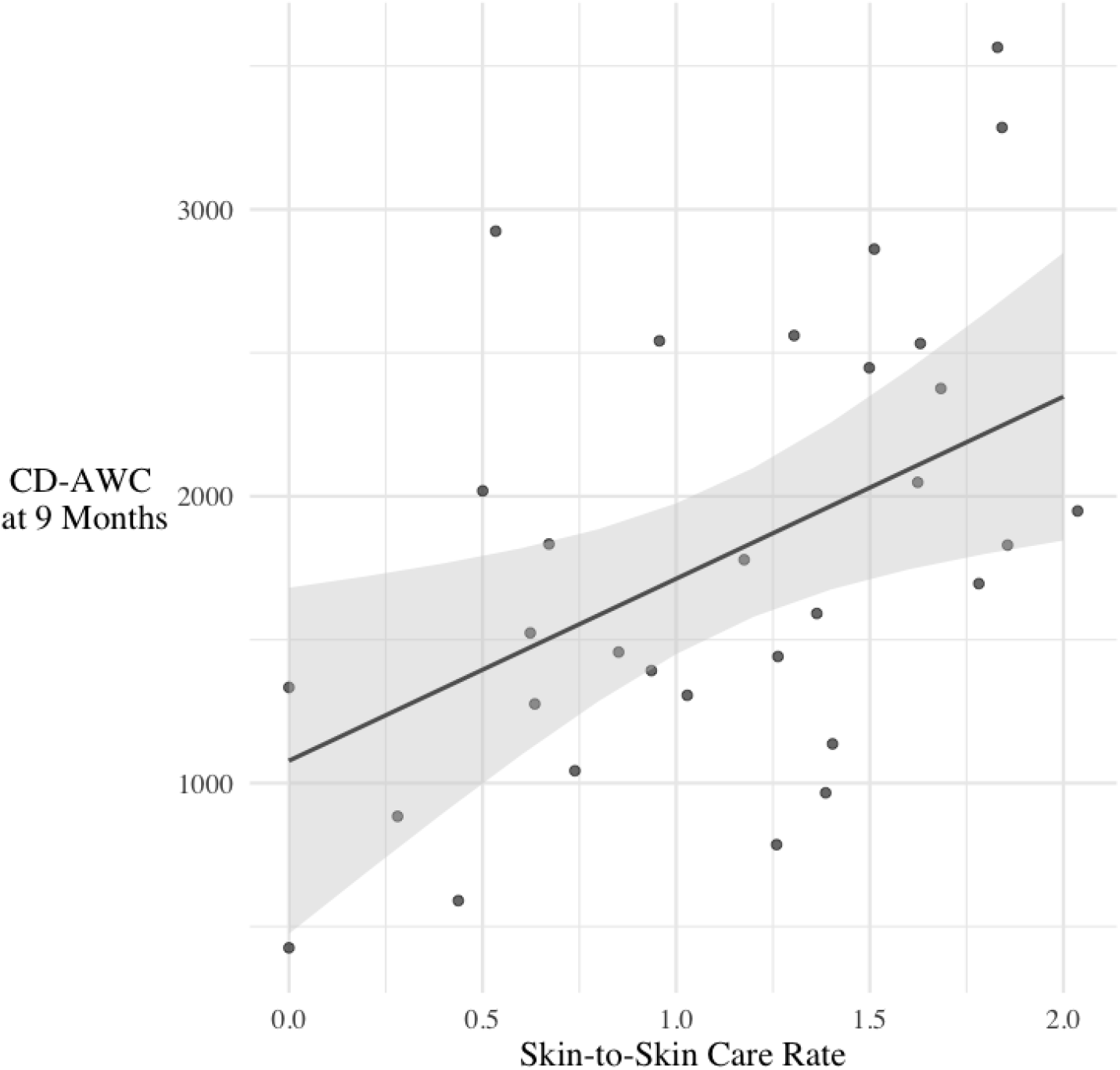
Model predictions showing relations between skin-to-skin (STS) rate in the NICU and child-directed adult word count (CD-AWC/hour) during day-long recording at 9 months (adjusted for degree of prematurity), controlling for covariates.

## Discussion

To investigate continuity in caregiver engagement for VPT infants, this study explored relations between STS care rates in the NICU and later caregiver verbal engagement. Our findings extend prior work^3,4^ by demonstrating that caregivers who engage in more STS care in the NICU are also more likely to verbally engage with their infant at home. Note that models controlled for visitation frequency, indicating that these effects were due to engagement in specific developmental care activities, rather than hospital attendance more generally. These findings reflect continuity in caregiver engagement from hospital to home.

There are many possible interpretations of the link between STS care rates and verbal engagement. STS care in early infancy increases oxytocin levels and strengthens parent-infant bonding, which may, in turn, enhance caregiver confidence and promote behaviors that support language-rich home environments.^9, 10^ Engaging in STS care may indirectly impact caregiver behaviors by reducing infant stress and promoting infant development, resulting in a relatively responsive child, which may lead to more responsive caregiving behaviors.^11,12^ Alternatively, engagement in STS care in the NICU may signal an underlying pattern of caregiver engagement that persists across time and caregiving contexts. In any case, clinical attention to the rates of STS care in the NICU may help to identify caregivers who might benefit from education and support in establishing favorable home environments for their VPT infants.

This study had both strengths and limitations. While our sample was diverse, it was relatively small, which may impact generalizability. Our measures of STS care relied on the accuracy of nurse charting. However, this practice is well-established at our center and integrated into standard clinical training.^13^ Moreover, STS care is a highly salient activity that involves active nurse participation, making it likely to be consistently and accurately recorded. Day-long audio recordings provide more objective, extensive, and less obtrusive estimates of caregiver verbal engagement than other available methods. However, they do not capture non-verbal behaviors that may also reflect positive caregiver-child engagement. Additionally, although caregivers were instructed to record a typical day at home, we could not verify who contributed to our measure of CD-AWC/hour. Finally, our design is correlational and therefore, we cannot draw causal conclusions from our findings.

In conclusion, this brief report highlights continuity in the caregiving environments of VPT infants at-risk for neurodevelopmental delays and underscores links between early developmental care and later language environments. Future work should examine the contributions of STS care and other developmental care practices in the NICU (e.g., verbal interactions) to caregiver verbal engagement in the home and subsequent neurodevelopmental outcomes. Routine charting of STS care offers a practical way for clinicians to monitor caregiver engagement in the NICU and to identify families who may benefit from additional post-discharge support. Promoting STS care as part of standard NICU practice may help establish enriched caregiving environments that support long-term developmental trajectories.

## Data Availability

Deidentified individual participant data will be made available upon request.

